# Type-2 immune skewing in patients with disseminated coccidioidomycosis

**DOI:** 10.1101/2025.09.26.25336729

**Authors:** Timothy J. Thauland, Smriti S. Nagarajan, Alexis V. Stephens, Samantha L. Jensen, Anviksha Srivastava, Miguel A. Moreno Lastre, Terrie S. Ahn, Chantana Bun, Michael T. Trump, Royce H. Johnson, George R. Thompson, Maria I. Garcia-Lloret, Valerie A. Arboleda, Manish J. Butte

## Abstract

**Background:** Disseminated coccidioidomycosis (DCM) is an often fatal and otherwise intractable condition requiring lifelong antifungal treatment. We have previously shown that a deranged polarization of CD4^+^ T cells toward a Type-2 phenotype can exist in the context of DCM. Here we studied a large population of subjects to determine the frequency of abnormal Type-2 skewing of CD4^+^ T cells in patients with coccidioidomycosis and to identify underlying genetic mechanisms supporting this skew.

**Methods:** We collected peripheral blood mononuclear cells from 204 patients with coccidioidomycosis, including 96 patients with disseminated disease. We measured immune phenotypes and cytokine production by CD4^+^ T cells from patients and healthy controls, and comparisons between groups were made based on disease severity and demographics. Whole genome sequencing was conducted on 149 individuals who also had cytokine profiling.

**Results:** We found that ~20% of DCM patients had a CD4^+^ T-cell compartment that was abnormally skewed toward a Type-2 (IFN-γ-IL-4+) phenotype. Type-2 skewing was highly correlated with male sex, with 80% of moderately skewed (Th2:Th1 ratio > 1.5) and 100% of severely skewed (Th2:Th1 ratio > 2) patients being male. Co-culture of T cells with the IL4R/IL13R-blocking antibody dupilumab rectified their Th1/Th2 skewing. Sequencing revealed rare variants in genes involved in the IL-12-IFN-γ axis in several Type-2 skewed patients, and we validated one such variant in *IFNGR1* as hypomorphic.

**Conclusion:** Patients with DCM, especially those who are male, should be screened for Type-2 skewing of CD4^+^ T cells. Patients with Type-2 skewing should be additionally screened for genetic defects in the IL-12-IFN-γ axis. Our findings give a mechanistic rationale for exploring blockade of IL4R as a treatment option in Type-2 skewed patients with refractory coccidioidomycosis.

## Introduction

Coccidioides fungi (*Coccidioides immitis* and *Coccidioides posadasii*) are endemic to arid regions of the American southwest and are responsible for a significant disease burden. There are ~200,000-350,000 infections per year in the United States, with 20,000 cases requiring medical attention and reaching clinical diagnosis (1). Approximately 60% of infections are asymptomatic, while in 40% of cases, coccidioidomycosis causes a relatively mild, self-limited respiratory disease (called “Valley fever”) (2). Approximately 1% of *Coccidioides* infections escape the lungs, resulting in disseminated coccidioidomycosis (DCM). DCM can manifest in the skin, bones, and nervous system causing significant morbidity and a mortality rate of 30% in the first 5 years after dissemination (3). If they survive, patients with DCM typically require antifungal treatment for life.

Known risk factors for progressing to DCM include age, sex, pregnancy, and race (4). Several monogenic variants in immune-related genes have been shown to predispose for DCM, including in the pathways recognizing β-glucan in the fungal cell wall or producing hydrogen peroxide in response to infection (5). Furthermore, patients with inborn defects in the IL-12/IFN-γ axis are at higher risk of developing DCM upon coccidioides infection (6–10). At present, there is no known laboratory test nor clinical algorithm available to predict whether infection with *Coccidioides* will remain as uncomplicated Valley fever or progress to dissemination.

Protective immunity to coccidioidomycosis requires a robust Type-1 response, characterized by the production of IFN-γ by *Coccidioides*-specific CD4^+^ Th1 cells (11). On the other hand, Type-2 immunity, characterized by the production of IL-4 and IL-13, confers susceptibility to invasive fungal infections by impairing innate immune defenses (12, 13). *Coccidioides* vaccines in development require IFN-γ production for protection (14–16). Recombinant IFN-γ has been successfully used to treat severe and refractory cases of coccidioidomycosis (17). Mechanistically, lung immune responses in mouse models that distinguish resistant strains (DBA/2) from susceptible ones (C57Bl/6) show that increased IFN-γ production correlates with resistance while increased IL-4 production correlates with susceptibility (18). Inducing Type-2 responses in the lung may be a form of immune evasion for certain fungi, driven by chitins in the fungal cell wall that elicit epithelial defenses including Type-2 alarmins (12), among other factors. In humans, elevations of IgE and eosinophilia have been associated with more severe coccidioidomycosis (19–21). Additionally, STAT3-mutated hyper-IgE (Job) syndrome, a disorder that skews T cells towards Type-2 immunity, has been reported in patients with severe coccidioidomycosis (22, 23).

The Pirofski-Casadevall damage-response framework suggests that therapeutic dampening of dysregulated inflammatory responses can be beneficial to the host (24). Indeed, targeted immunosuppression can over the detrimental effects of Type-2 immunity: antibody blockade of IL-4 in mouse models reduces fungal burden (25). We previously reported a case of life-threatening DCM characterized by a defect in Type-1 immunity and who also showed severe Type-2 skewing (26). We found that the patient’s CD4^+^ T-cell blasts showed a strong Type-2 phenotype, with a large majority of cytokine-producing T cells making IL-4 but not IFN-γ. The patient was successfully treated with a combination of IFN-γ (augmenting Type-1 responses) and an anti-IL-4R monoclonal antibody (dupilumab, mitigating Type-2 responses). These treatments markedly improved the patient’s lesions concomitant with an increase in IFN-γ producing Th1 cells and a decrease in IL-4 producing Th2 cells (26). This case offered hope that a subset of severe coccidioidomycosis patients could benefit from an immunomodulatory strategy.

In most healthy people, CD4^+^ T cells that are restimulated ex vivo are skewed towards a Type-I phenotype (27). Our success in treating a DCM patient with augmented Type-2 skewing coupled with the knowledge that Type-1 immunity is essential for a successful immune response to coccidioides infection led us to ask how prevalent the Type-2 phenotype is in patients with clinically significant coccidioidomycosis.

## Results

Our patient cohort were drawn from the central valley of California and surrounding counties. They were recruited either at the Valley Fever Institute in Bakersfield or the Valley Fever Center of Excellence at the University of California, Davis. Our patient cohort was divided into the following clinical categories that we have previously described: 2: uncomplicated coccidioidomycosis; 3: complicated pulmonary disease (including patients with fibro-cavitary disease, persistent disease despite six months of treatment, and respiratory failure); 4: disseminated disease without meningitis; 5: disseminated disease with meningitis (28). The relevant characteristics of our patient cohort are shown in **Table 1**. Of note, approximately two-thirds of individuals with complicated pulmonary and disseminated disease were male, and over half of our cohort self-identified as Hispanic or Latino. These demographics are in line with previous population-based studies on coccidioidomycosis disease burden in the central valley of California (29). The design of our study is shown in **Figure 1**.

**Figure 1.**
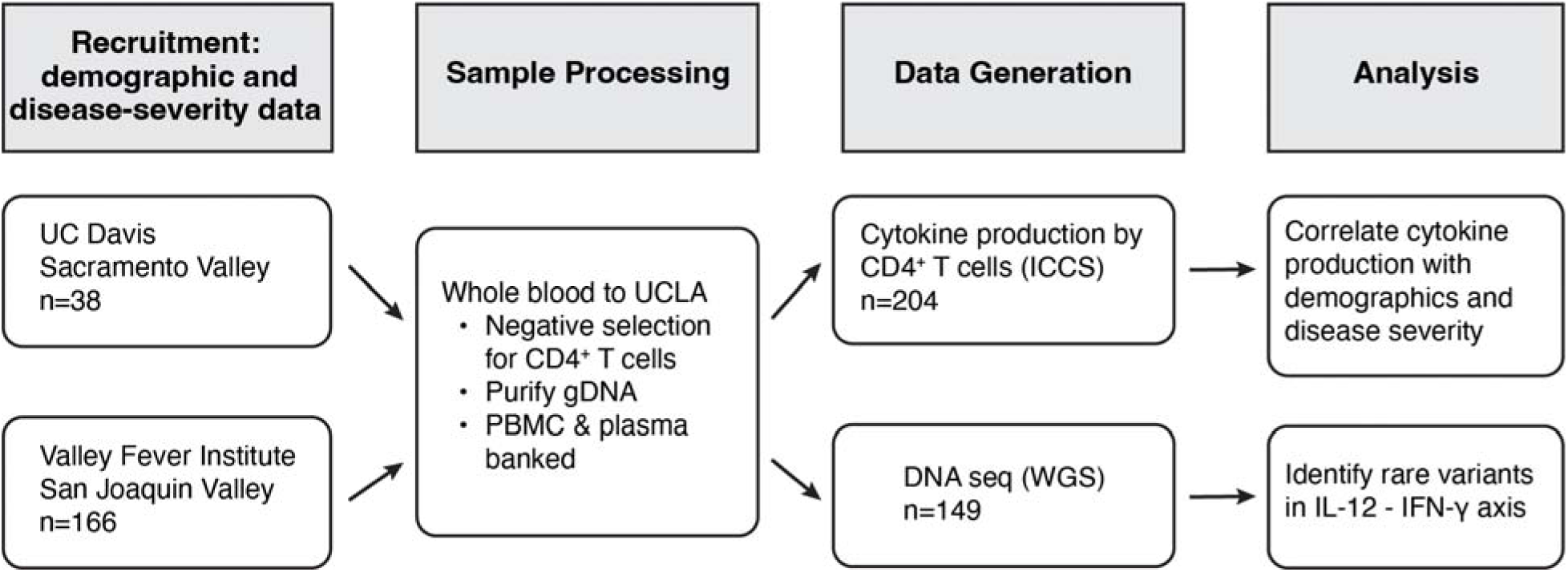
Flowchart demonstrating experimental design from recruitment to analysis.

**Table 1.** Patient characteristics.

|  | **Coccidioidomycosis Clinical Category** |  |  |  |
| --- | --- | --- | --- | --- |
|  | **2: Pulmonary**<br>**n=66 (32%)** | **3: Complicated**<br>**Pulmonary**<br>**n=42 (21%)** | **4: Disseminated**<br>**n=38 (19%)** | **5: Disseminated**<br>**with meningitis**<br>**n=58 (28%)** |
| **Age (y) <sup>A</sup>** | 18-76 (41) | 23-74 (46) | 23-82 (50) | 14-75 (46) |
| **Sex** |  |  |  |  |
| Female | 36 (55%) | 14 (33%) | 9 (24%) | 20 (34%) |
| Male | 30 (45%) | 28 (67%) | 29 (76%) | 38 (66%) |
| **Self-identified Race & Ethnicity** |  |  |  |  |
| Asian | 4 (57%) | 2 (29%) | 1 (14%) | 0 |
| Black | 3 (25%) | 4 (33%) | 2 (17%) | 3 (25%) |
| Hispanic or Latino | 30 (31%) | 20 (21%) | 15 (16%) | 31 (32%) |
| White/Hispanic or Latino | 5 (36%) | 3 (21%) | 2 (14%) | 4 (29%) |
| White | 17 (33%) | 10 (19%) | 14 (27%) | 11 (21%) |
| Other/More than one race | 3 (33%) | 2 (22%) | 3 (33%) | 1 (11%) |
| Unreported | 4 (29%) | 1 (7%) | 1 (7%) | 8 (57%) |
| **Recruitment location** |  |  |  |  |
| UC Davis | 11 (29%) | 8 (21%) | 8 (21%) | 11 (29%) |
| Valley Fever Institute | 55 (33%) | 34 (20%) | 30 (18%) | 47 (28%) |
| <sup>A</sup>Range (Median) |  |  |  |  |

To measure cytokine production, we purified CD4^+^ T cells from both patients and healthy controls, activated them polyclonally, and performed intracellular cytokine signaling (ICCS) on acutely restimulated cells. The proportions of Th1 cells (defined as IFN-γ^+^ IL-4^−^) and Th2 cells (defined as IL-4^+^ IFN-γ^−^) were determined (**Figure 2A**) and the Th2:Th1 ratio was calculated, ignoring those cells that expressed both cytokines. We established a cutoff point for abnormal Type-2 skewing at a Th2:Th1 ratio of 1.5. All healthy control subjects (n=48) fell below this cutoff point, save one (Th2:Th1 ratio = 1.52). The proportion of coccidioidomycosis patients with a Th2:Th1 ratio greater than 1.5 increased with increasing disease severity, with significantly more DCM patients with a Type-2 skewed phenotype than controls or patients with pulmonary-only disease (**Figure 2B** and **Figure S1A**). Seventeen percent of DCM patients were Type-2 skewed, with many having more than a 2:1 ratio of Th2:Th1 cells. These results reveal that a large fraction of patients with severe coccidioidomycosis including DCM showed Type-2 skewing.

**Figure 2.**
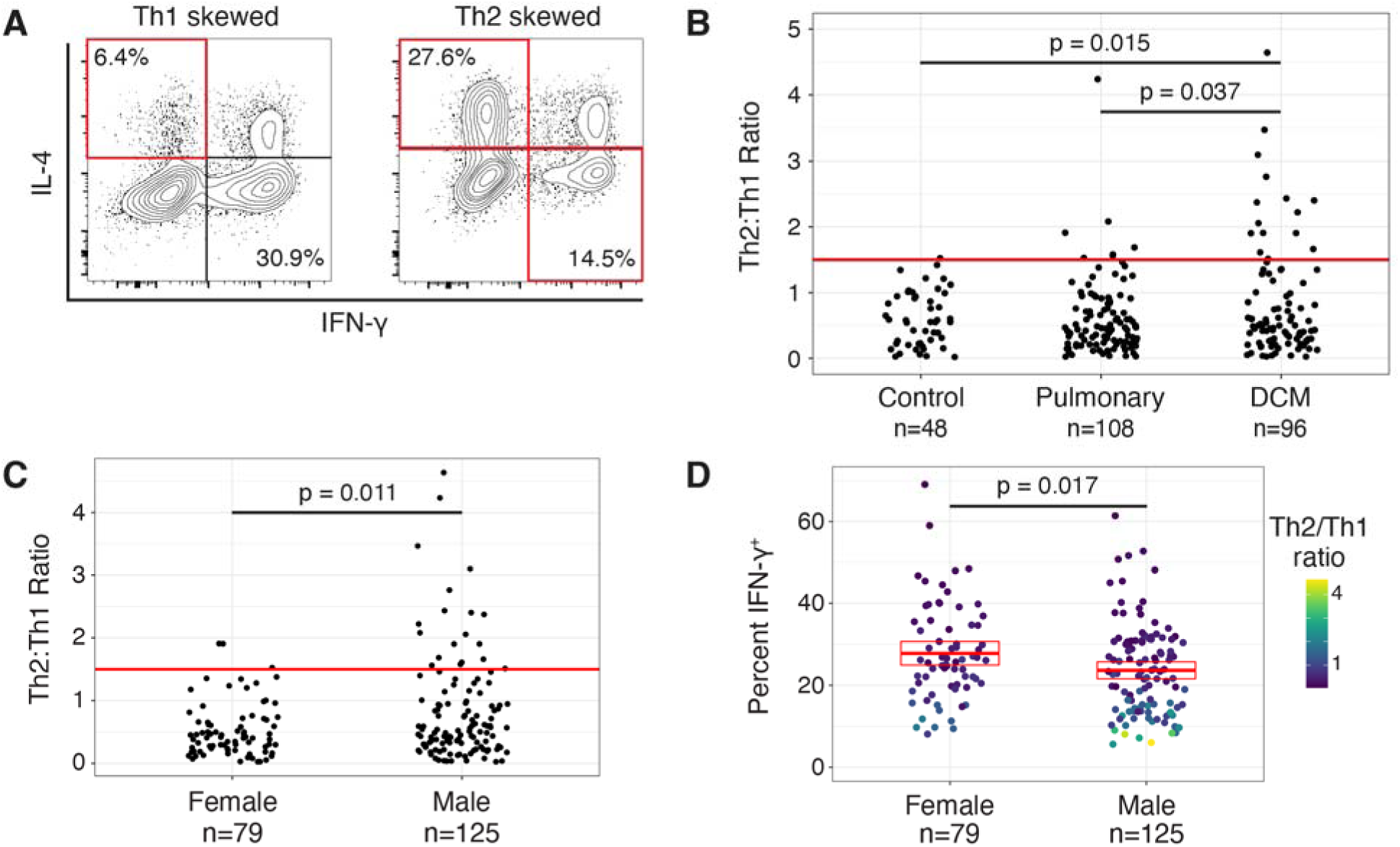
Type-2 skewing of CD4+ T cells in coccidioidomycosis. **(A)** ICCS data for expression of IFN-γ and IL-4 was used to generate Th2:Th1 ratios. An example of a Type-1 and Type-2 skewed patient is shown. **(B)** Type-2 skewing in controls and patients with pulmonary disease or DCM. Type-2 skewing is significantly more common in DCM patients. (Barnard’s unconditional test) **(C)** Th2:Th1 ratio stratification by sex reveals that Type-2 skewed individuals are predominantly male. (Barnard’s unconditional test) **(D)** Females had more IFN-γ producing cells than males on average. This difference was driven entirely by the subset of males with a high Th2:Th1 ratio. The means (horizontal line) and 95% CI (box) are shown. (Wilcoxon’s rank sum test). Each dot is one subject.

There are conflicting data whether there is an inherent difference in Th1/Th2 bias in males and females (30, 31), with some studies showing enhanced Type-1 skewing in males (32) and others in females (33). Given that male sex is a risk factor for progression to DCM (34), we stratified our patient cohort by sex and examined the Th2:Th1 ratio. We found a strong bias towards male sex in Type-2 skewed patients (**Figure 2C**). Approximately 80% of patients with Th2:Th1 ratio greater than 1.5, and 100% of patients with a ratio greater than 2, were male. Females on average made significantly more IFN-γ than males (**Figure 2D**). These differences were driven entirely by the presence of patients with a Th2:Th1 ratio greater than 1.5, as removal of these individuals from the statistical analysis yielded no significant sex-related differences in IFN-γ or IL-4. (IFN-γ: [Male]=26.2%, [Female]=28.6%, p=0.17; IL-4: [Male]=10.6%, [Female]=10.0%, p=0.52). These results demonstrated an unexpected and significant sex bias of Type-2 skewing in patients with severe coccidioidomycosis.

Some studies have shown that age may affect Th1/Th2 balance, with older adults biased towards Type-2 responses, although contradictory results also exist (35–37). When we stratified our patient cohort by age and examined cytokine production, there was no discernible pattern. Type-2 skewed patients were found in every age cohort (**Figure S1B**). These results show no relationship between Type-2 skewing and age.

Patterns of chemokine receptor expression on memory CD4^+^ T cells have been used as surrogate markers for helper T-cell polarization (38). Ex vivo cytokine analysis is laborious, so we measured chemokine receptor expression on CD4^+^ memory T cells from our patient cohort to determine if these surrogate markers could be used to discern subjects who showed Type-2 skewing (**Figure S2A**). We compared the percent of CD4^+^CD45RO^+^ memory cells with a CCR6^−^ CXCR3^+^CCR4- “Th1” or CCR6-CXCR3^−^CCR4^+^ “Th2” phenotype against the measured Th2:Th1 ratio of cytokine production (**Fig S2B-C**). The relationship between chemokine receptor expression and cytokine production showed the expected slope (i.e.. a negative slope when percent cells with a “Th1” chemokine receptor phenotype is plotted against Th2:Th1 ratio). However, only the correlation of “Th2” memory cell levels with Th2:Th1 ratio reached statistical significance, and the contribution of percent “Th2” memory cells to the variance in Th2:Th1 ratio was modest (R^2^ = 0.12). These results suggest that chemokine receptor expression on CD4+ T cells is not a good surrogate for cytokine production in these subjects.

Type-2 skewing of immune responses is found in atopic disease, in many cases characterized by eosinophilia and elevated levels of serum IgE (39). We sought to understand if eosinophilia was seen in those subjects with Type-2 skewing in our coccidioidomycosis cohort. We compared eosinophil counts or total serum IgE at the time of blood collection with Th2:Th1 ratio, but no clear trends were present (**Figure S3A** and **Figure S3B**). We note that not all patients had laboratory findings available, and only a handful of patients in our entire cohort had either eosinophilia or elevated IgE. Additionally, there was no correlation between eosinophil counts and IgE levels among patients for whom both measurements were available (**Figure S3C**). These results show that neither eosinophilia nor IgE levels are predictive of Type-2 skewing in our coccidioidomycosis cohort.

IL-17 production by Th17 cells is crucial for mucosal defenses to yeast and other fungal infections, and both vaccine studies in mice (40, 41) and ex vivo analysis of T cells from pediatric patients (42) have suggested that Th17 cells offer protection from coccidioidomycosis. Crucially, there is evidence that Th17 cells are important for preventing progression to DCM (4). Thus, we examined IL-17A production by CD4^+^ T cell blasts (**Figure 3A**). Compared to healthy controls, we found increased IL-17A production in coccidioidomycosis patients. However, we did not observe any differences in Th17 skewing between uncomplicated and DCM patients (**Figure 3B**). These results show that while increased Th17 skewing is associated with coccidioidomycosis, the degree of skewing does not correlate with the severity of disease. When patients were segregated by sex, we did find lower IL-17A production in male patients, and this difference was driven by patients with a Th2:Th1 ratio greater than 1.5 (**Figure 3C**). This result indicates that Type-2 skewing may come at the expense of Th17 defenses in some male subjects.

**Figure 3.**
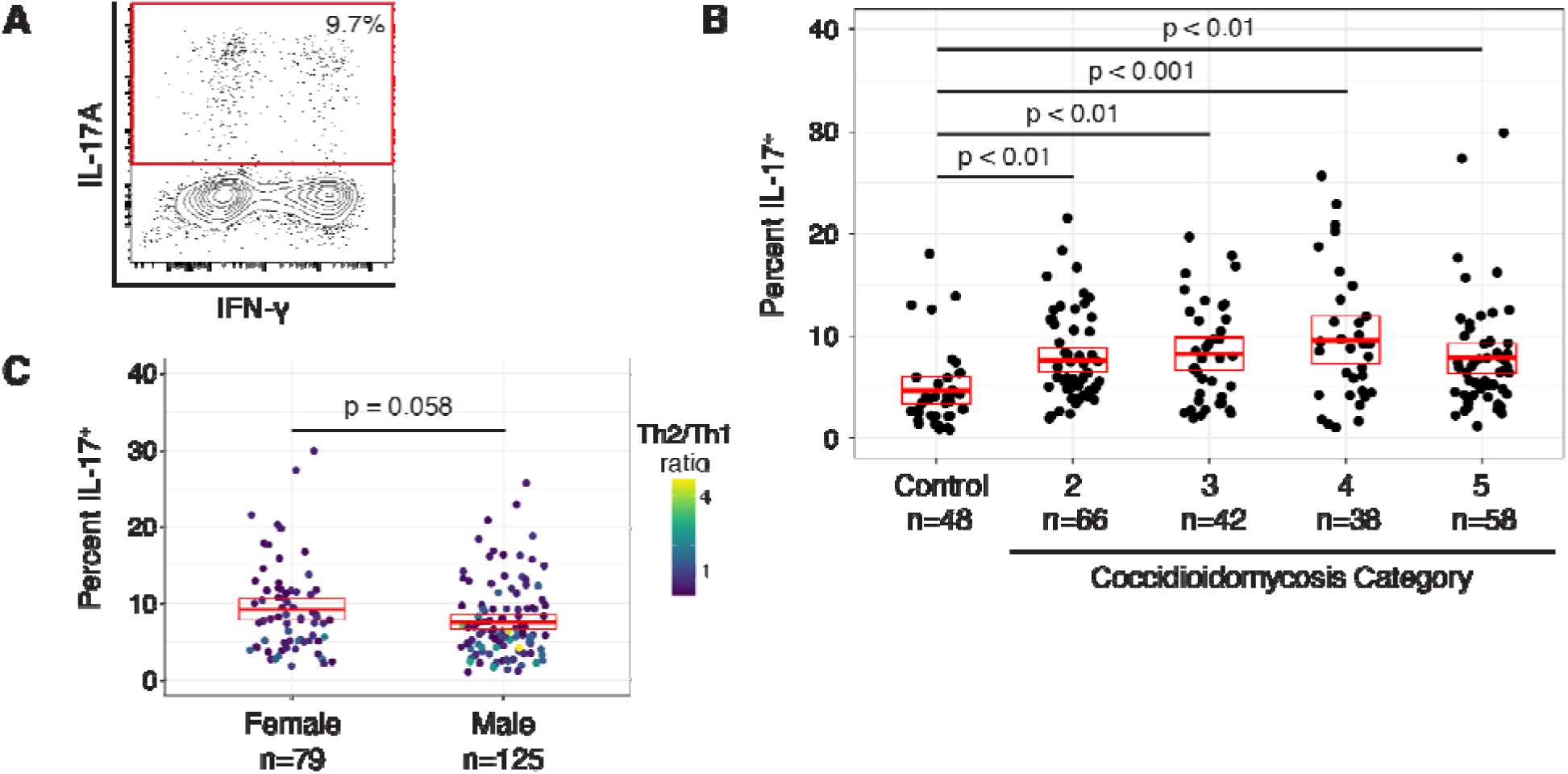
Th17 polarization is associated with coccidioidomycosis but not severity of disease. **(A)** Example of ICCS for measurement of Th17 polarization. **(B)** Coccidioidomycosis patients have significantly more Th17 cells than healthy controls, but no differences are seen when patients are stratified by disease severity. (Kruskal-Wallis rank sum test with Dunn’s post-hoc test and Bonferroni correction) **(C)** Male patients had fewer IL-17A producing cells on average than females. (Wilcoxon’s rank sum test). The means (horizontal line) and 95% CI (box) are shown. (Wilcoxon’s rank sum test). Each dot is one subject.

We have previously demonstrated the efficacy of treating a patient with refractory DCM and Type-2 skewing with IFN-γ and dupilumab (IL-4R blockade) (26). To demonstrate the efficacy of dupilumab alone in remodeling cytokine production by CD4^+^ T cells ex vivo in patients with coccidioidomycosis, we cultured CD4^+^ T cells from patients in our cohort with or without dupilumab and measured cytokine production by ICCS (**Figure 4**). Four of these patients had a Th2:Th1 ratio greater than 1.5. T cell blast cultures from all patients showed an increase in the percentage of IFN-γ producing cells, and 12 out of 13 showed a decrease in IL-4 producing cells and Th2:Th1 ratio in the presence of dupilumab (**Figure 4B-D**). These ex vivo results support the notion that dupilumab may be able to remodel *Coccidioides*-specific memory CD4^+^ T cells in patients. Indeed, we have subsequently treated several Type-2 skewed coccidioidomycosis patients with dupilumab and demonstrated an alteration of cytokine production ex vivo— toward a Type-1 phenotype – and clinical improvement (manuscript in preparation).

**Figure 4.**
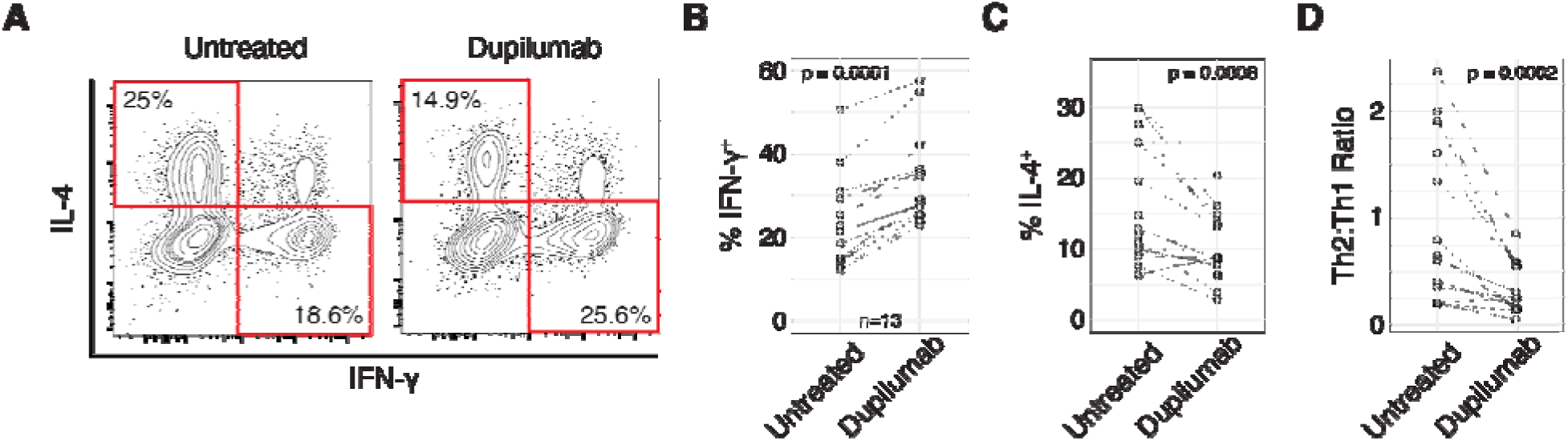
Dupilumab restores Type-1 responses in subjects with Type-2 T cell skewing. **(A)** Representative examples of cytokine production by CD4^+^ T cells cultured in vitro with or without dupilumab. **(B-D)** Effect of dupilumab treatment on percent IFN-_γ_^+^ cells **(B)**, percent IL-4^+^ cells **(C)**, or Th2:Th1 ratio **(D)**. (Wilcoxon’s signed-rank exact test)

Patients with defects in Type-1 immunity are known to be susceptible to severe coccidioidomycosis (6). To determine if deleterious alleles in the IL-12-IFN-γ axis were enriched in our cohort of Type-2 skewed patients, we employed whole genome sequencing. Both genomic data and cytokine profiling were collected for 149 patients. We identified 74 rare protein-coding variants in genes in the IL-12-IFN-γ axis that were carried by at least one patient with a Type-2 phenotype. Six rare missense variants were found only in DCM patients with a Type-2 phenotype (**Table 2**). One, *IL12RB1* G447A, occurs with an allele frequency of 3.7 × 10^−6^ in gnomAD v4 and is predicted to be pathogenic by CADD, SIFT, PolyPhen, and ESM1b.

**Table 2.** Rare inborn variants in the IL-12-IFN-_γ_ axis found only in Type-2 skewed subjects.

| Protein Variants |  |  |  | Patient Characteristics |  |  |  |
| --- | --- | --- | --- | --- | --- | --- | --- |
| Gene | Variant | Allele Freq. | CADD Score | Category | Sex | Age | Th2:Th1 Ratio |
| IL12RB1 | G447A | 3.73E-06 | 23.0 | 4 | M | 38 | 1.28 |
| IFNGR1 | P431L | 3.78E-05 | 13.8 | 5 | F | 59 | 1.91 |
| IFNGR2 | F16FA | 2.56E-05 | 10.4 | 5 | M | 63 | 3.47 |
| IL12RB1 | A91T | 8.47E-04 | 0.79 | 3 | M | 57 | 1.40 |
| IFNGR2 | S81Y | 6.20E-07 | 0.56 | 5 | M | 14 | 1.46 |
| IL12RB2 | I41V | 3.05E-05 | 0.35 | 5 | M | 63 | 1.51 |

Another, *IFNGR1* P431L, occurs with an allele frequency of 3.8 × 10^−5^ and is listed as a variant of uncertain significance (VUS) in the ClinVar database for disseminated atypical mycobacterial infection. We tested both rare variants in functional assays of cytokine signaling. Both B cells and monocytes from the patient with a P431L variant in *IFNGR1* had a strikingly hypomorphic response in the phosphorylation of STAT1 upon stimulation with IFN-γ (**Figure 5**). Cells from the patient with a G447A variant in *IL12RB1,* however, responded normally to IL-12 stimulation as measured by phosphorylation of STAT4 (**Figure S4**). These results demonstrate the feasibility of identifying rare mutations in signaling pathways critical for Type-1 immunity in patients with Type-2 skewed CD4^+^ T cells.

**Figure 5.**
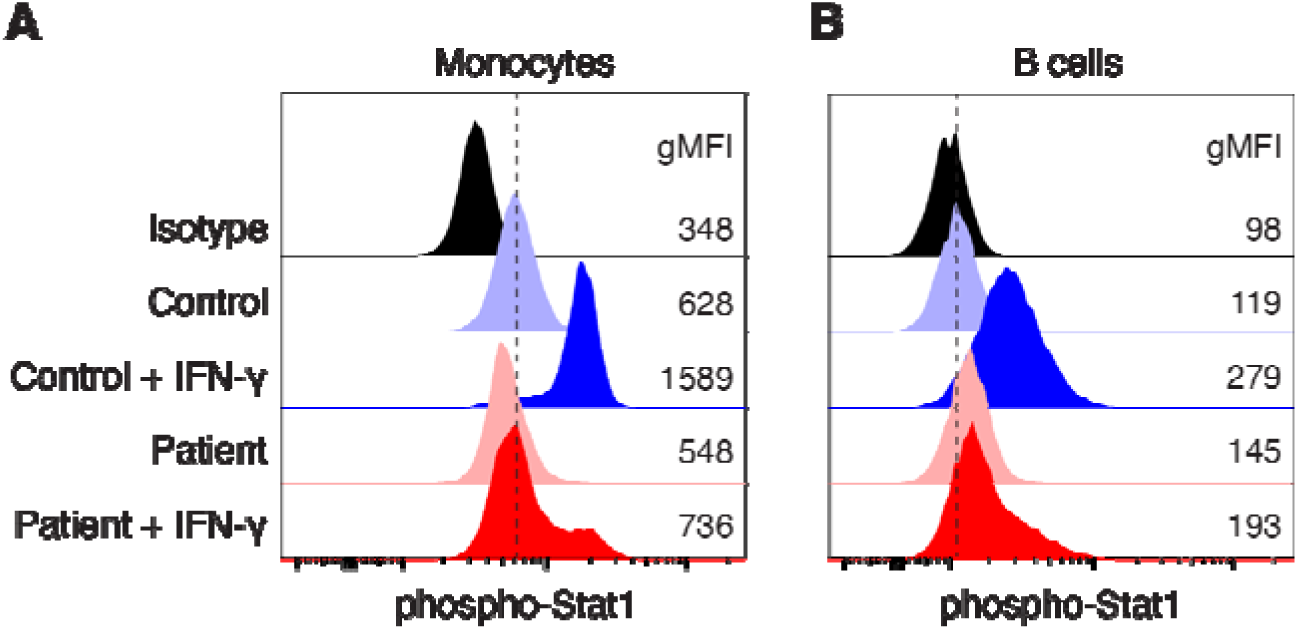
Validation of a rare variant in *IFNGR1* in a Type-2 skewed subject with DCM. A heterozygous P431L variant in IFNGR1 in a Type-2 skewed coccidioidomycosis patient (“Patient”) results in a hypomorphic response to IFN-_γ_. **(A and B)** Measurement of phosphorylation of Stat1 in response to IFN-_γ_ stimulation shows a defective response in B cells **(A)** and monocytes **(B)** from a patient with a P431L mutation in *IFNGR1* and a healthy control subject. The geometric mean fluorescence intensities for all peaks are shown.

## Discussion

This work sheds light on how dysregulated immune responses may leads to severe coccidioidomycosis. We have demonstrated three important new findings: (1) Type-2 skewing of CD4^+^ T cells is present in a significant fraction (~20%) of patients with DCM. This phenotype is correlated with disease severity: significantly more Type-2 skewed individuals are found in the patient cohort with disseminated disease, especially in patients with nervous system involvement (Category 5). (2) Males are much more likely to have Type-2 skewed T cells than females, and Type-2 skewing in these individuals is concomitant with a decrease in Th17 cells. These results are concordant with the long-standing observation that males are more susceptible to severe coccidioidomycosis. It is noteworthy that most male patients with disseminated disease were not Type-2 skewed, and thus the overrepresentation of males among patients with severe disease is multi-factorial and may include non-biological factors (e.g., overrepresentation of males in certain occupations, including construction and farm work). Nevertheless, our data show that screening male patients with severe disease for Type-2 skewing will be fruitful, given the availability of pharmaceutical interventions (IFN-γ and anti-IL-4R) for this population. (3) Genetic defects in the IL-12-IFN-γ signaling axis are known to dramatically increase susceptibility to mycobacterial infections (Mendelian susceptibility to mycobacterial disease) (43), and have been implicated in several patients with severe coccidioidomycosis (6). Here, we show that rare variants in the IL-12-IFN-γ axis exist in Type-2 skewed patients with DCM. As a proof of principle, we measured signaling through the IFN-γ receptor in a patient harboring a heterozygous mutation in the IFNGR1 gene and showed a hypomorphic response.

One limitation of our study is the use of bulk CD4^+^ T-cell cultures to measure cytokine production. Our rationale for using bulk cultures is two-fold: 1) Cytokine production by bulk CD4^+^ T cells cultured under neutral conditions will necessarily be determined by a unique combination of genetics and epigenetic programming (determined by previous infections) that is reflective of an individual’s predisposition toward Type-1 or Type-2 T-cell responses. 2) In a previous report, we showed that cytokine production in bulk T cells closely correlates with the T-cell response to *Coccidioides* antigens ex vivo (26), and we have subsequently treated multiple Type-2 skewed patients with IFN-γ and/or dupilumab with encouraging results (manuscript in preparation). Ideally, cytokine production by T cells specific for *Coccidioides* antigens would also be measured. Unfortunately, in our hands, there are no existing coccidioides antigenic reagents that reproducibly activate T cells. For example, the T27 *Coccidioides* antigenic preparation (44, 45) stimulates robust upregulation of activation induced markers (CD69, CD134, and CD137) or cytokine production in fewer than half of clinically-proven coccidioidomycosis patients. Our group and others (46) are currently developing *Coccidioides*-derived antigenic preparations to find reliable reagents for stimulating T cells in coccidioidomycosis patients with diverse HLA haplotypes.

These data are of actionable clinical significance due to the demonstrated importance of Type-2 immunity in mediating dysregulated responses to *Coccidioides* infection. We have demonstrated clinical success in blocking Type-2 immunity with an anti-IL-4R monoclonal. We suggest that patients with persistent *Coccidioides* infections, especially those with disseminated disease and those not responding to antifungals, and especially males, should have their Th2:Th1 ratios tested ex vivo. We also suggest the actionability of screening patients found to have Type-2 skewing for functionally relevant genetic defects in the IL-12-IFN-γ axis. Knowledge of such defects could inform treatment decisions and potentially improve patient outcomes.

## Methods

### Ethics Approval

All patients provided informed consent to participate in protocols approved by the Institutional Review Board (IRB) of the University of California, Los Angeles, with reliance agreements from the Valley Fever Institute and the University of California, Davis.

### Patient Recruitment

All patients were evaluated by an infectious disease specialist from the Valley Fever Institute or the University of California, Davis, and were confirmed to have coccidioidomycosis. These experts also reviewed each patient’s disease trajectory and clinical status to assign scores using a clinicopathological categorization system that we have previously published (28).

Patients were subsequently enrolled in an IRB-approved protocol, under which whole blood was collected and shipped to UCLA for analysis.

### Sex as a biological variable

Patients were recruited without bias regarding biological sex. In the final cohort used in this report, there were 79 females and 125 males. The excess of males in our study is in line with previous reports demonstrating that severe coccidioidomycosis is more common in males (4). Healthy controls were anonymous, and neither their biological sex nor age is known.

### T cell culture

CD4^+^ T cells were purified from fresh or frozen PBMCs by EasySep negative selection (StemCell, Cat #17952). T cell blasts were generated by plating ~10^6^ purified CD4^+^ cells on 12 well plates coated with 1 μg/mL anti-CD3 (clone OKT3; Biolegend) in complete T cell medium (RPMI 1640 supplemented with 10% FCS, 10 mM HEPES, 1 mM sodium pyruvate, 55 μM 2-mercaptoethanol, and 1X Pen-Strep) with 2 μg/mL soluble anti-CD28 (clone CD28.2; Biolegend). On day 2 after stimulation, cells were replated in fresh medium supplemented with 50 U/mL IL-2. Cytokine production was measured on day 7-8. In some experiments, 10 μg/mL anti-IL-4R monoclonal antibody (dupilumab) was included in the T cells cultures for the entire duration of their culture.

### Intracellular cytokine staining

Approximately 10^6^ CD4^+^ T cell blasts were acutely stimulated with 50 ng/mL PMA and 1 μM ionomycin for 5 h and 1x GolgiPlug (BD; Cat#555029) was added for the final 4 h of the stimulation. Cells were then fixed with PBS / 2% PFA and permeabilized with 1x Perm Buffer (Biolegend, Cat #421002). Cells were stained in Perm Buffer with anti-IFN-γ (Biolegend; clone 4S.B3), anti-IL-4 (Biolegend; clone MP4-25D2), and anti-IL-17A (Biolegend; clone BL168). Data were collected on a Cytek DxP10 digital flow cytometer and analyzed with FlowJo software.

### Phospho-Stat Assay

Thawed PBMCs were stimulated with 10 ng/mL rhIFN-γ (PeproTech, Cat# 300-02) or rhIL-12 (PeproTech, Cat# 200-12) for 20 min at 37 °C. Stimulation was stopped by adding an equal volume of pre-warmed Cytofix buffer (BD, Cat# 554655) and incubating for 12 min at 37°C. Fc receptors were blocked (TruStain FcX; Biolegend) for 5 min at RT, followed by a 20 min stain on ice for CD19 (clone HIB19; Biolegend) and CD14 (clone HCD14; Biolegend) or CD4 (clone RPA-T4; Biolegend). Cells were permeabilized for 30 min on ice with 1 mL pre-chilled Phosflow Perm Buffer III (BD, Cat# 558050). After permeabilization, the cells were stained with anti-pStat1 pY701 (clone 4a; BD), anti-pStat4 p693 (clone 38/p-Stat4; BD) or isotype control (MOPC-173; BD) for 25 min at RT. Data were collected on a Cytek DxP10 flow cytometer and analyzed with FlowJo software.

### DNA sequencing and imputation

We conducted whole genome sequencing (WGS) from whole blood for 156 patients from our cohort. For the first batch of 88 samples, DNA was isolated using a Qiagen blood extraction kit, prepared with an Illumina TruSeq DNA PCR-free library kit, and sequenced using an Illumina NovaSeq6000. The DRAGEN Germline Pipeline v3.2.8 was used to align and map reads to the hg38-alt-aware reference on Illumina BaseSpace. Small variant joint calling was carried out with the DRAGEN Joint Genotyping Pipeline v3.7.5. The second batch of 68 participants was sequenced with Illumina Novaseq X Plus and processed using a similar DRAGEN pipeline on Amazon Web Services (AWS) designed by the UCLA SwabSeq lab. PIPELINE was used to align and map reads to the hg38_alt_aware genome build.

The two WGS batches of gVCFs were merged and indexed by chromosome using bcftools (7, 8). The WES imputed VCFs were then merged with the WGS chromosome-level gVCFs and indexed with the same method. Variant INFO field annotations were recalculated in the merged files with the bcftools +fill-tags plug-in and alternate alleles not appearing in any sample were removed to convert to the final merged VCFs. The chromosome-level VCFs were then merged.

### Quality control of sequencing data

The final merged VCF underwent quality control (QC) using bcftools (47, 48). Variants that had any VCF filter other than “PASS” were removed. The genotype for any sample who did not have a quality score of more than 20 or total read depth of more than 10 was set to missing. Duplicate variants were removed. We produced variant depth metrics, as well as sample missing genotype and singleton counts, to use for further QC. We then converted the VCF to PLINK format and standardized variant naming. We filtered out variants with missing genotypes for more than 10% of samples, variants with a Hardy-Weinberg Equilibrium p-value < 0.0001, and samples with more than 10% of variants with missing genotypes (49–51). We used PLINK 2 to calculate KING-robust estimates of sample relatedness and removed one of each pair of individuals with a kinship coefficient > 0.177 to remove first-degree relationships (52, 53). We also checked that the genetic sex of each patient matched the reported sex in our records. After QC, we had sequencing data for 149 patients with calculated Th2:Th1 ratios.

### Risk gene variant annotation and pathogenicity prediction

From merged VCF files, we filtered to include only those variants falling in one of nine genes involved in Th cell differentiation and signaling: *IL12A*, *IL12B*, *IL12RB1*, *IL12RB2*, *IFNG*, *IFNGR1*, *IFNGR2*, *ISG15*, and *TYK2*. We then used the Ensembl Variant Effect Predictor (VEP) to predict the consequences of each variant and add pathogenicity annotations (54). We also assessed the potential pathogenicity of all missense variants using the ESM1b and AlphaMissense machine learning models (55, 56). In R, we identified which variants passed quality control and calculated their allele frequencies in just the QC-passed samples by coccidioidomycosis type (UVF, CPC, and DCM) and by status of Type-2 skewing (skewed, not skewed). Focusing on the variants with protein-coding consequences in their Ensembl canonical transcripts, we filtered to include only those variants that had at least one carrier with Type-2 status assessed. We selected rare variants (gnomAD AF < 0.01) with a higher allele frequency in DCM patients than in patients without dissemination (UVF and CPC) and in Type-2 skewed patients than in those without Type-2 skew. Finally, we manually selected variants to prioritize for functional validation based on CADD Phred score, ClinVar annotations, and missense severity scores.

### Statistical Analysis

When comparing categorical data, we used Barnard’s unconditional two-sided test. When comparing the means of groups, we used the Shapiro-Wilk test to determine if the data were normally distributed. For comparisons where data from at least one group was not normally distributed, we used non-parametric tests: the Kruskal-Wallis rank sum test and Dunn’s post-hoc test with Bonferroni correction for multiple comparisons, and the Wilcoxon rank sum test with continuity correction for comparison of two groups. For experiments with paired samples, we used the Wilcoxon signed-rank exact test. Linear regressions and R^2^ values were generated in R with the function geom_smooth and method = lm. The linear relationship between two variables was tested with Pearson’s product-moment correlation test. All statistical analyses were conducted in R.

## Data Availability

All data produced in the present work are contained in the manuscript.

## Data Availability

Values for all data points in graphs are reported in the Supporting Data Values file.

## Author Contributions

Conceptualization: TJT, MIG-L, MJB

Investigation: TJT, SSN, AVS, TSA, MAML, SLJ, CB, MST

Visualization: TJT, SSN, SLJ

Funding acquisition: MJB

Supervision and project administration: MJB, MIG-L, VAA

Writing – original draft: TJT

Writing – review & editing: everyone

## Data Availability

All data produced in the present work are contained in the manuscript.

## Acknowledgements

We acknowledge support for sequencing from the UCLA SwabSeq lab.

