## Supplemental for "Type-2 immune skewing in patients with disseminated coccidioidomycosis" (from another version of this preprint: version 1, 2025-09-28)

### Supplemental Figures

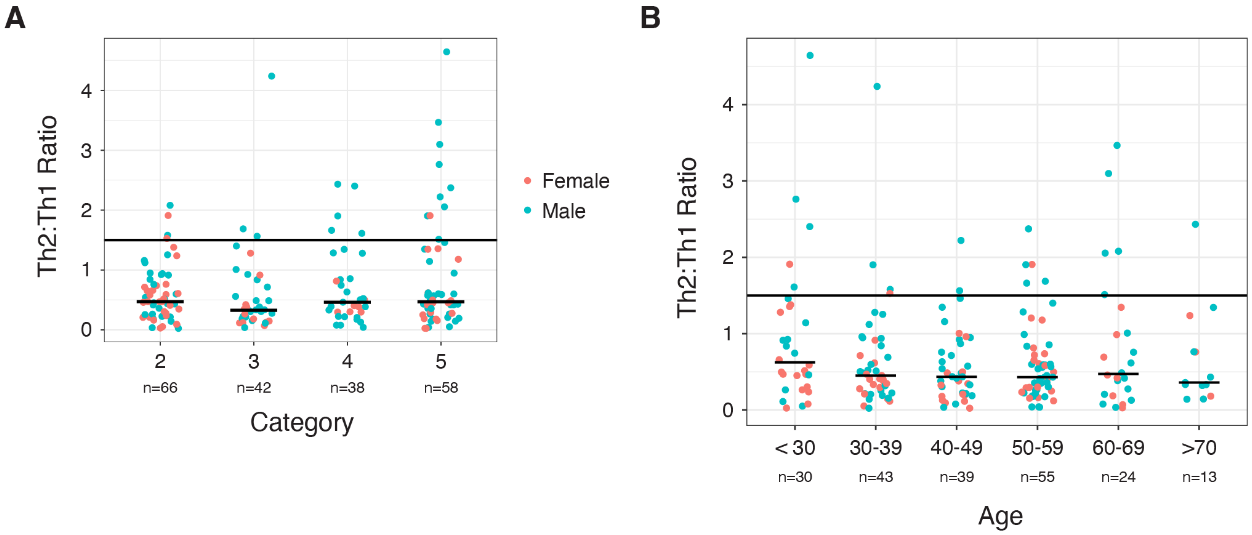


**Supplemental Figure 1. (A)** Distribution of Type-2 skewed individuals across the four clinical categories. **(B)** Type-2 skewing in coccidioidomycosis patients is not correlated with age. Medians are shown.

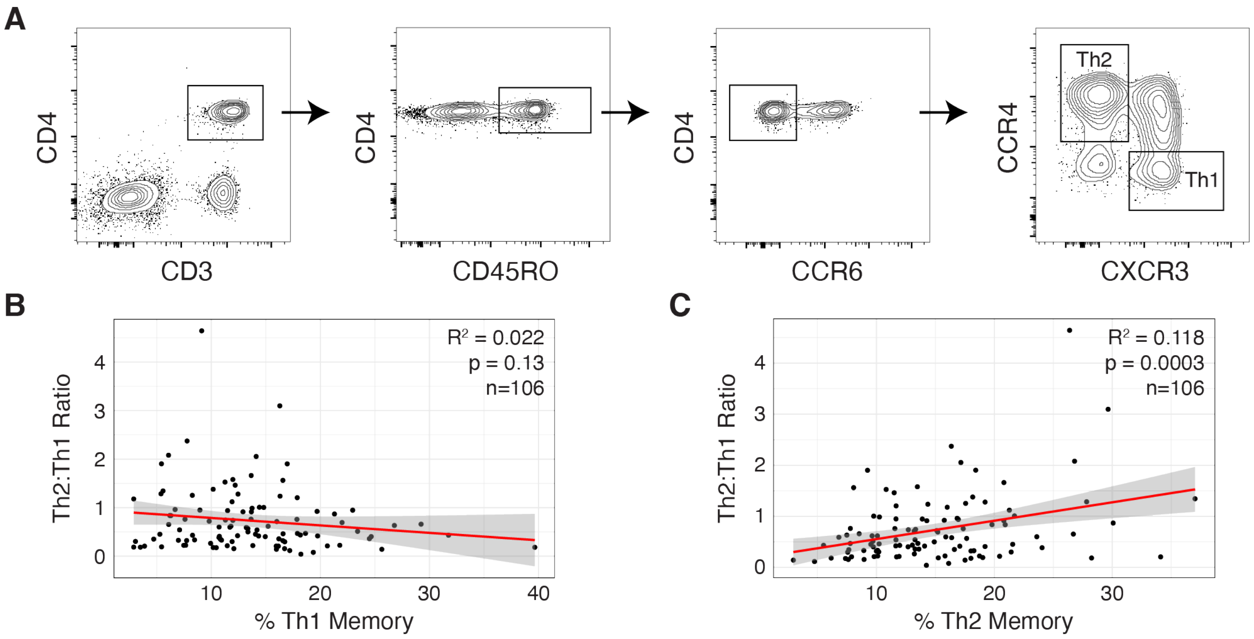


**Supplemental Figure 2. (A)** Gating scheme for identifying memory Th1 and Th2 cells. **(B and C)** Linear regressions with 95% CI between the Th2:Th1 ratio and the percent Th1 ($CCR6^{-}CXCR3^{+}CCR4^{-}$) **(B)** or Th2 ($CCR6^{-}CXCR3^{-}CCR4+$) **(C)** cells among $CD4^{+}CD45RO^{+}$ memory T cells in coccidioidomycosis patients. (Pearson's product-moment correlation test)

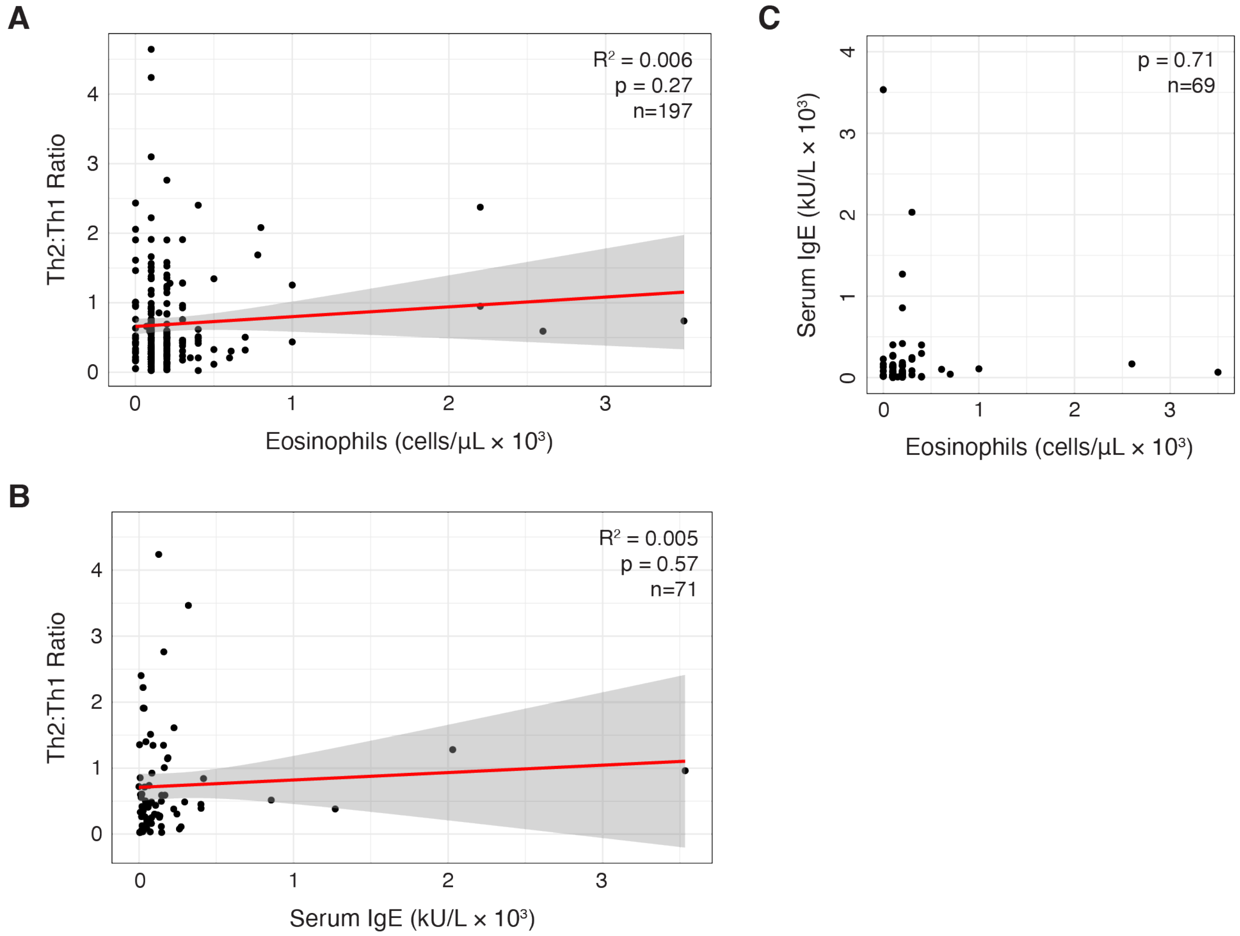


**Supplemental Figure 3. Chemokine receptor expression and cytokine production by CD4+ T cells. (A and B)** Linear regressions with 95% CI between the Th2:Th1 ratio and absolute eosinophil counts (A) or serum IgE (kU/L) (B). (Pearson's product-moment correlation test) **(C)** Absolute eosinophil counts are plotted vs serum IgE, showing the absence of a relationship in our dataset.

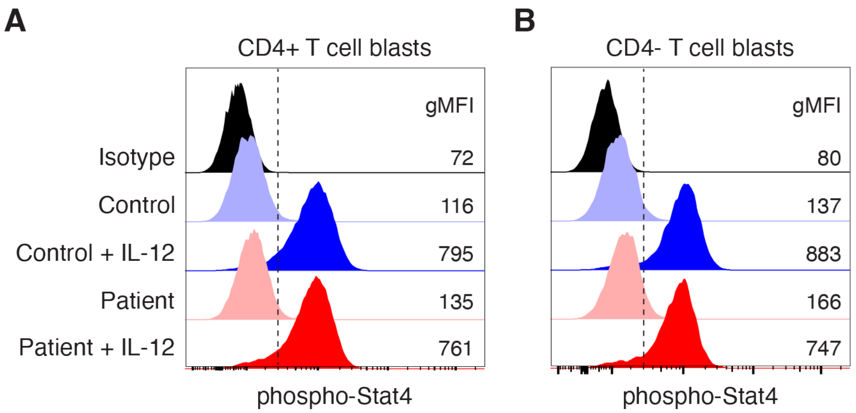


**Supplemental Figure 4. (A and B)** Measurement of phosphorylation of Stat4 in response to IL-12 stimulation shows a normal response in CD4$^{+}$ **(A)** and CD4$^{-}$ **(B)** T cell blasts from a patient with a G447A mutation in *IL12RB1*. The geometric mean fluorescence intensities for all peaks are shown.
